# NEURAL RESPONSE TO THREAT AND REWARD AMONG YOUNG ADULTS AT RISK FOR ALCOHOL USE DISORDER

**DOI:** 10.1101/2023.07.20.23292969

**Authors:** Katelyn T. Kirk-Provencher, Rosa H. Hakimi, Keinada Andereas, Anne E. Penner, Joshua L. Gowin

## Abstract

Alcohol use disorder is 50% heritable; those with positive family histories represent an at-risk group within which we can test anticipation of threat and reward prior to development of harmful alcohol use. We examined neural correlates of the interaction between family history, threat anticipation (unpredictable threat), and monetary reward anticipation, in a sample of healthy young adults with (*n*=31) and without (*n*=44) family histories of harmful alcohol use. We used a modified Monetary Incentive Delay task with sustained threat of hearing a scream during fMRI. We examined the interaction between family history group, anticipation of threat, and anticipation of reward in the insula, nucleus accumbens, and medial prefrontal cortex. Family history positive individuals showed less activation in the left insula during both safe and threat blocks compared to family history negative individuals (*p*=0.005), but the groups did not differ as a function of unpredictable threat (*p*>0.70). We found an interaction (*p*=0.048) between cue and group in the right nucleus accumbens where the family history positive group showed less differentiation to the anticipation of gaining $5 and losing $5 relative to gaining $0. The family history positive group also reported less excitement for trials to gain $5 relative to gaining $0 (*p*<0.001). Prior to chronic heavy alcohol use, individuals with, relative to without, enriched risk may have diminished reward processing via both neural and behavioral markers to potential rewarding and negative consequences. Neural response to unpredictable threat may not be a contributing factor to risk at this stage.

## Introduction

Alcohol use contributes to 3.8% of global deaths (1) and alcohol use disorder is approximately 50% heritable (2, 3). This enriched risk via genetic predisposition presents a challenge for distinguishing risk patterns at the individual level, and an opportunity to study biological markers that may indicate risk for developing harmful patterns of alcohol use among currently healthy young adults. Some individuals who develop an alcohol use disorder may engage in harmful and hazardous use due to rewarding effects of alcohol, including the reduction in anxious states (4, 5) and negative affect (6–8) which can be the result of threatening stimuli. However, it remains unclear whether alteration in processing threat and reward are present prior to developing an alcohol use disorder. Therefore, examining the effect of family history of harmful alcohol use on the anticipation of sustained threat and anticipation and receipt of monetary reward, may help us better understand whether these constructs are risk factors for developing an alcohol use disorder.

Greater harmful patterns of alcohol use (5), family history of alcohol use disorder, and diagnosis of alcohol use disorder (9) are associated with increased anticipatory anxiety to unpredictable threat. Unpredictable threat results in increased insula activation among individuals with alcohol use disorder (10). Additionally, in healthy adults, threat of aversive stimuli (e.g., exposure to unpleasant images or pain) results in altered neural circuitry in reward and loss processing regions (11–13). For example, healthy adults with greater self-reported stress showed less activation in the medial prefrontal cortex after receiving feedback for both winning and losing money (13).

More research has examined the effect of family history of harmful alcohol use on the anticipation of reward using the Monetary Incentive Delay task (14–19), while fewer studies have examined family history in relation to outcomes of receiving or losing monetary reward (e.g., 14, 17). For example, altered reward processing has been demonstrated in children, adolescents, and young adults with family history of alcohol use disorder, such that those with positive family histories relative to those with negative family histories demonstrate decreased activation in the nucleus accumbens during the anticipation of reward (16, 20) and loss (19), while family history positive individuals show no significant difference in activation during the outcome of reward (14).

While previous research has separately examined reward and threat processing in the context of alcohol use disorder risk, it is possible that neural response to aversive, unpredictable threat may modulate response to possible rewards, such as monetary rewards. Those with family history of harmful alcohol use may be less sensitive to threat, more sensitive to reward, or an interaction of both. However, to our knowledge, no studies have examined the possibility of anticipation of threat modulating anticipation of reward in an enriched risk sample of currently healthy young adults.

The goal of this study is to identify neural markers of risk in young adults who have not yet transitioned to heavy alcohol use, particularly focusing on neural response to an induced anticipatory anxiety to threat and anticipation of monetary reward. While neural response to reward anticipation has been well-characterized using the Monetary Incentive Delay (MID) task (see 21, 22-25), our study is the first to use the addition of sustained threat of a scream (MID-Scream; see 26) to examine the interaction of anticipation of threat and anticipation of reward in family history positive and negative young adults. We aimed to test the main and interaction effects of family history, anticipation of threat, and anticipation of reward to better understand the ways in which these factors may contribute additive or multiplicative risk for developing an alcohol use disorder. We hypothesized that during the MID-Scream task, the family history positive group will sustain higher activation levels in the insula and medial prefrontal cortex during the anticipation of threat, while the family history negative group will have dampened neural response in these regions. Further, the family history positive group versus the family history negative group will have less reactivity in the nucleus accumbens during the anticipation of a possible monetary reward.

## Materials and Methods

### Procedures

Participants were recruited through advertisements for a study (approved by the Colorado Multiple Institutional Review Board) examining neural correlates of parental and sibling harmful alcohol use. Participants provided informed consent. They reported their current age, sex, race, ethnicity, and alcohol consumption behaviors. Participants were administered a semi-structured interview to assess psychiatric diagnoses by trained clinicians (Structured Clinical Interview for DSM-5 Disorders Reserach Version; 27). We created dichotomous variables for both past and current alcohol use disorder and cannabis use disorder. Data were collected from March 2019 through June 2021 and stored using REDCap (28).

Eligibility criteria were (1) being able to undergo MRI, (2) aged 18-22 (one individual turned 23 prior to the MRI scan), (3) had used alcohol (4) scored less than seven on the Alcohol Use Disorder Identification Test (indicating non-hazardous drinking; 29), (5) not using medications affecting the hemodynamic response, (6) not seeking treatment for an alcohol use disorder, (7) not being treated for a psychiatric disorder, (8) scored less than 11 on the Cannabis Use Disorder Identification Test-Revised (30) or less than eight uses of cannabis per month, (9) not using tobacco regularly (< 20 cigarettes per week), (10) less than 10 lifetime uses of illicit drugs, (11) no prescription medication misuse in the past 12 months, (12) no past head trauma with unconsciousness lasting more than 10 minutes, and (13) not pregnant. At the beginning of the study visit, participants completed a second MRI safety screener to ensure MRI eligibility, an alcohol breathalyzer, saliva drug screener, and females took a urine pregnancy test.

### Measures

#### MID-Scream Task

During the MRI scan, participants completed the MID-Scream task that was designed to examine anticipation of threat (e.g., sustained anticipation of aversive stimuli) modulated neural response to potential monetary rewards; full details of this task are described elsewhere (26). Briefly, during the anticipation phase of the task, participants were shown one of three cues indicating the opportunity to gain $5, avoid losing $5, or gain $0 (no change in earnings). The screen had a colored border indicating whether the trial was in the safe condition (yellow) or threat condition (blue), where the threat condition indicates the possibility of hearing a scream and seeing a scary face. After being presented with the cue and condition combination, participants were briefly shown a target, and were instructed to press a button as quickly as possible. During the outcome phase of the task, participants were provided feedback on their responses to the target during each trial. Successful responding during the circle cue results in gaining $5, while unsuccessful responding during the triangle cue results in losing $5. Success or failure during the square cue results in no change in earnings (i.e., gain $0); see Figure 1. A trial includes the anticipation plus outcome phase. Safe and threat conditions occurred in blocks of nine consecutive trials. There were six blocks per run. There were two runs of the task. Thus, there were 18 trials total of each of the six combinations (gain $5-threat, gain $5-safe, lose $5-threat, lose $5-safe, gain $0-threat, gain $0-safe). There was an additional block during which participants heard the scream and saw the scary face once during each of the two runs (i.e., saw the face and heard the scream twice during the course of the task). This block was removed from analyses a priori as this portion of the task design differs from the trials of interest and due to concern about movement in the scanner following the aversive stimuli. Each run lasted 13 minutes. Participants were informed that they would receive their earnings during the MID-Scream task prior to entering the MRI machine, and were paid their earnings at the end of the study.

**Figure 1.**
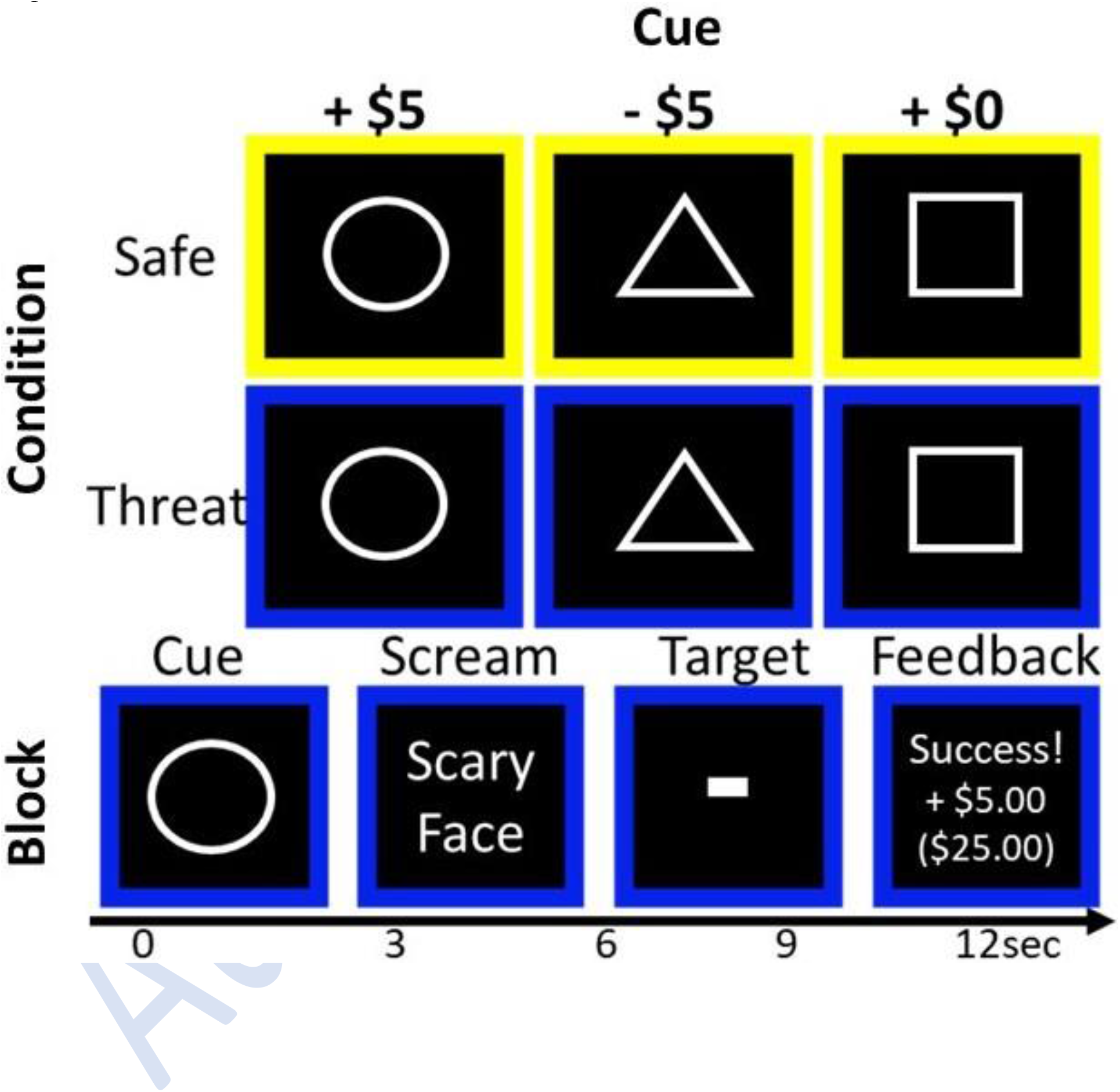
This figure depicts a schematic of the MID-Scream Task. This is an example of a Threat Block. During this trial of the threat block, the participant was presented with the possibility to gain $5 (circle cue) during sustained threat of hearing a scream and seeing a scary face (blue condition) during the anticipation phase of the task. Upon successfully pressing the button while the target was shown, the participant was provided feedback that they were successful, had gained $5, and shown their cumulative earnings, during the outcome phase of the task.

#### Self-Report Ratings

Following the MRI scan, participants were asked to report how they felt in response to each cue and condition combination, and to hearing the scream during the MID-Scream task. Using a scale of 0-10, they reported how much they liked each combination, and how excited and nervous each combination made them feel.

#### Family History of Harmful Alcohol Use

Using the Family History Assessment Module (31), participants reported whether first-degree relatives (i.e., parents and siblings) had experienced legal, occupational, or social problems due to their alcohol use, given that parental and sibling alcohol use is associated with alcohol use risk (e.g., 32, 33, 34). Those with first-degree relatives with harmful alcohol use only (*n* = 11) or both harmful alcohol and other drug use (*n* = 20) were coded as family history positive. All others were coded as family history negative (*n* = 44).

#### Additional Measures

Depression was assessed using the Beck Depression Inventory (35), a 21-item measure assessing the intensity of depression. Anxiety was assessed with the Spielberger State-Trait Anxiety Inventory (36), a 40-item self-report measure assessing state anxiety (anxiety taking place at a given time) on 20 items, and trait anxiety (the relatively stable disposition to anxiety) on 20 items.

### Imaging Acquisition

Magnetic resonance imaging scans were conducted using a Siemens 3.0 Tesla Skyra magnet with a 20-channel head coil. Functional images were acquired using BOLD signal across 40 axial slices with TR = 2000ms, TE = 30ms, flip angle = 77°, acquisition matrix = 74mm x 74mm voxels, 40 axial 3mm thick/0mm gap slices, and multiband factor of 2.

### fMRI Data Preprocessing

Imaging data were preprocessed using Analysis of Functional NeuroImages version 22.2.01 (AFNI; 37). We first converted raw scanner data into AFNI compatible formats. Using @SSwarper, anatomical data were nonlinearly warped to the Montreal Neurological Institute (MNI) standard space and were skull-stripped and deobliqued. Task data were deobliqued to match anatomical data. Time points greater than 0.3mm Euclidean distance of framewise displacement were censored from analyses. We also censored timepoints with greater than 10% of voxels displaying outliers in activation levels. We used an 8mm kernel for blur. Each run was scaled to produce a mean voxel intensity of 100. For regression, we examined six cue conditions (gain $5-threat, gain $5-safe, lose-$5-threat, lose-$5-safe, gain$0-threat, gain$0-safe) and 12 outcome conditions (trial success and failure for each of the six cue conditions). We used a hemodynamic response model with 1-second block and an amplitude of 1 for each of these 18 conditions. For the main effect of threat block, we applied a 96-second block model with amplitude of one, starting when the screen indicated if the participant was about to begin a safe or threat block, and terminating when the block was over. For each of those regression analyses, we included six regressors of no interest to control for motion of translation and rotation in the *x*, *y*, and *z* dimensions. Final voxel resolution was 3mm isotropic.

### Data Analytic Plan

Data analyses were conducted in *R* version 4.1.2.We assessed significant differences between family history positive and family history negative groups on demographic, substance use, depression, and anxiety variables, and behavioral task data. We used Student and Welch two-sample *t*-tests and Wilcoxon rank sum tests for continuous variables, and Fisher’s Exact Test and Pearson Chi-Square tests for categorical variables. Linear mixed-effects models and post hoc comparisons using factorial analysis of variance (ANOVA) with Kenward-Roger approximation assessed the main and interaction effects of family history group, cue, and condition on participant MID-Scream reaction times and success rates, and self-report ratings following the task.

#### Regions of Interest (ROI) Analyses

Left and right nucleus accumbens masks were defined using the Harvard-Oxford Subcortical Structural Atlas (https://identifiers.org/neurovault.image:1700), and the left and right medial prefrontal cortex masks were defined by creating 8mm diameter spheres centered on the MNI coordinates of x = ± 6, y = 49, z = –8 in AFNI (38). The insula masks were defined using the Brainnetome Atlas BN_Atlas_246_3mm.nii (39) using parcels 167 and 168 for the left and right, respectively. Values were extracted from each mask using AFNI’s *3dROIstats* command. For the bilateral nucleus accumbens, we assessed the main effects of group, cue, and condition during the anticipation phase, as well as interaction effects of group-by-cue, group-by-condition, and group-by-cue-by-condition. In the bilateral medial prefrontal cortex, we assessed the same effects during the outcome phase of the task. In the bilateral insula, we assessed the main effects of group and block (safe and threatening blocks of the task), and we assessed the interaction effects of group-by-block.

Analyses were conducted using the *nlme* package in *R* software. The dependent variable in analyses were the beta weights for each task effect. Group, cue, and condition were entered as fixed-effects in restricted maximum likelihood linear mixed effects models, with participants entered as the random effect variable. The models were assessed in *R* followed by factorial ANOVA with Kenward-Roger approximation. Post hoc comparisons of least-square means with Tukey adjusted *p*-values were conducted using the *lsmeans* package in *R* to assess the significant main and interaction effects. To account for the power to detect interactions being half of main effects, an alpha level of 0.10 was applied to interactions. For the ROIs, sensitivity analyses were conducted to examine potential confounding factors of lifetime alcohol or cannabis use disorders, sex, depression, and trait anxiety by entering the covariates into adjusted linear mixed effects models.

#### Exploratory Analysis

To assess whether group differences in neural activation were evident in other brain regions, we conducted exploratory whole-brain analyses using a linear mixed effects model in AFNI. We used a minimum voxel-wise threshold of *p* < 0.001 and a family-wise corrected error of α = .05 as assessed by a Monte Carlo simulation using AFNI’s *3dClustSim* command with spatial autocorrelation function correction parameters of 0.57, 7.04, and 14.59. This indicated a minimum cluster size of *k* ≥ 49 voxels. For identified clusters, we examined effects in *R*. As neural activation during anticipation of reward and during sustained threat were our primary interests, we assessed the magnitude of effect of group during the gain $5 cue (reward anticipation) and during the threatening block. We generated separate effect size maps for reward and threat as a function of family history group using Cohen’s *d*, as recommended (40) for transparency in neuroimaging research.

## Results

### Participant Characteristics

Participants were aged 18-23, mostly female (56%) and White (76%). The family history positive group (*n* = 31) had a greater proportion of females (χ^2^(1) = 8.41, *p* < 0.05) and reported greater state and trait anxiety (both *t*_73_ > 3.0, *p* < 0.05) than the family history negative group (*n* = 44). The groups did not significantly differ on remaining characteristics or alcohol consumption behaviors (Table 1).

**Table 1.** Participant Characteristics and Substance Use by Family History Group.

| Variable | FH-<br>(*n* = 44) | FH+<br>(*n* = 31) | Test Statistic |
| --- | --- | --- | --- |
|  | *n* (%) |  |  |
| Sex |  |  |  |
| Male | 26 (59.09) | 7 (22.58) | $\chi^2(1) = 8.41$ |
| Female | 18 (40.91) | 24 (77.42) |  |
| Race |  |  |  |
| American Indian/Alaska Native | 0 (0.00) | 1 (3.23) | Fisher's *p* = .12 |
| Asian | 4 (9.09) | 1 (3.23) |  |
| Black/African American | 2 (4.55) | 1 (3.23) |  |
| White | 30 (68.18) | 27 (87.10) |  |
| Biracial | 6 (13.64) | 0 (0.00) |  |
| No Response/"Unknown" | 2 (4.54) | 1 (3.23) |  |
| Ethnicity |  |  |  |
| Hispanic | 11 (25.00) | 7 (22.58) | Fisher's *p* = 1.00 |
| Non-Hispanic | 32 (72.73) | 24 (77.42) |  |
| No Response/"Unknown" | 1 (2.27) | 0 (0.00) |  |
| Alcohol Use Disorder |  |  |  |
| Past | 1 (2.27) | 3 (9.68) | Fisher's *p* = .300 |
| Current | 0 (0.00) | 1 (3.23) | Fisher's *p* = .413 |
| Cannabis Use Disorder |  |  |  |
| Past | 1 (2.27) | 1 (3.23) | Fisher's *p* = 1.00 |
| Current | 1 (2.27) | 1 (3.23) | Fisher's *p* = 1.00 |
| High Motion in Scanner | 3 (4.00) | 2 (2.67) | Fisher's *p* = 1.00 |
|  | *M* (*SD*) |  | Test Statistic |
| Depression | 4.09 (4.63) | 6.45 (7.55) | *t*(46) = 1.55 |
| Anxiety |  |  |  |
| State Anxiety | 30.64 (7.94) | 37.35 (8.77) | *t*(73) = 3.46 |
| Trait Anxiety | 33.34 (8.79) | 41.10 (9.69) | *t*(73) = 3.61 |
| Current Age | 20.96 (1.37) | 21.02 (1.33) | *t*(73) = 0.20 |
| Alcohol Use Disorder Identification Test | 4.61 (2.28) | 4.55 (2.59) | *t*(73) = 0.12 |
|  | Median (IQR) |  | Test Statistic |
| Percent Days Drank Alcohol in Past Year <sup>a</sup> | 10 (13.75) | 7 (15.00) | *W* = 610.50 |
| Number of Binge Episodes in Past Year <sup>b</sup> | 2.50 (9.00) | 2.00 (4.50) | *W* = 708.00 |
| Total Drinks on Average Drinking Occasion <sup>c</sup> | 3 (2.00) | 3 (1.00) | *W* = 657.00 |
| Percent of Scan Data Censored (% TRs) <sup>d</sup> | 3.82 (6.91) | 4.12 (6.99) | *W* = 675.00 |

Note. FH+ = family history positive, FH- = family history negative, IQR = interquartile range.
<sup>a</sup> FH+ ranged from 1-75%, and FH- ranged from 0-60%.
<sup>b</sup> FH+ ranged from 0-50, FH- ranged from 0-50.
<sup>c</sup> FH+ ranged from 0-24, FH- ranged from 0-14.
<sup>d</sup> FH+ ranged from 0-21.76, and FH- ranged from 0-36.06%.
Bold font indicates significance at *p* < .05.

### Behavioral Outcomes of the MID-Scream Task

#### Reaction Time and Success Rate

Reaction time and success rate for trials indicated that participants were engaged and motivated by the task cues (see Supporting Information).

#### Self-Report Ratings

There were no significant group-by-cue-by-condition interactions for liking, excitement, or nervousness (all *F*_2,365_ < 0.20, *p* > 0.80; Table S1). There were significant main effects of condition on liking and nervousness (both *F*_2,365_ > 26.0, *p* < 0.001; see Table S1), such that participants reported liking the safe condition more than the threat condition and were more nervous during the threat condition than the safe condition.

There were significant group-by-cue interactions on liking, excitement, and nervousness (all *F*_2,365_ > 2.0, *p* < 0.070; Table S1); see Figure 2. Compared to the family history positive group, the family history negative group reported a greater relative difference in liking cues to gain $5 versus gain $0. The family history negative group was significantly more excited during cues to gain $5 than the family history positive group (*p* = 0.032), and also showed a greater relative change for excitement to gain $5 versus gain $0. Compared to the family history positive group, the family history negative group reported a greater relative difference in nervousness for cues to gain $5 and avoid losing $5 relative to cues to gain $0. See Supporting Information for full results of self-report ratings following the MID-Scream task.

**Figure 2.**
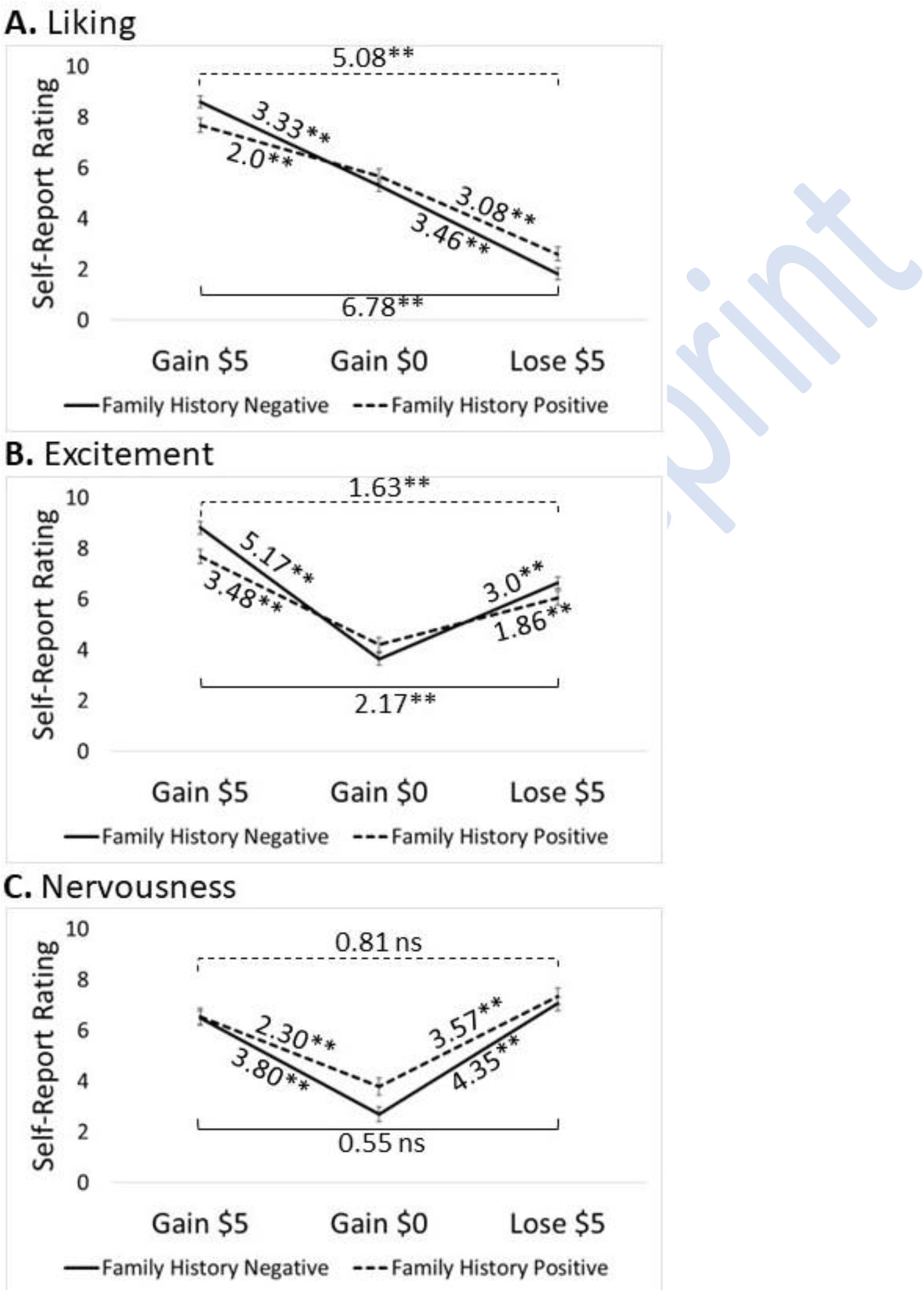
This figure depicts the results of self-report ratings linear mixed effects modeling. Values indicate the average ratings, ** indicates significance of p < 0.001, “ns” indicates non-significance, and all error bars represent the standard error of the least-squares means. **Panel A:** The graph depicts the significant group-by-cue interaction for liking (*F*_2,365_ = 6.86, *p* = 0.001) with significant contrasts. **Panel B:** The graph depicts the significant group-by-cue interaction for excitement (*F*_2,365_ = 8.93, *p* < 0.001) with significant contrasts. **Panel C:** The graph depicts the group-by-cue interaction for nervousness (*F*_2,365_ = 2.83, *p* = 0.060) with significant contrasts.

### Region of Interest Analyses

#### Insula

There were no significant group-by-block interactions in the left or right insula (see Table 2). There was a significant main effect of block on neural activation bilaterally in the insula (both *F*_1,73_ > 11.0, *p* = 0.001) where participants demonstrated greater activation during the sustained threat blocks relative to the safe blocks. In the left insula, there was a significant main effect of group (*F*_1,73_ = 8.51, *p* = 0.005) where the family history negative group showed greater neural activation during the task (Figure 3).

**Figure 3.**
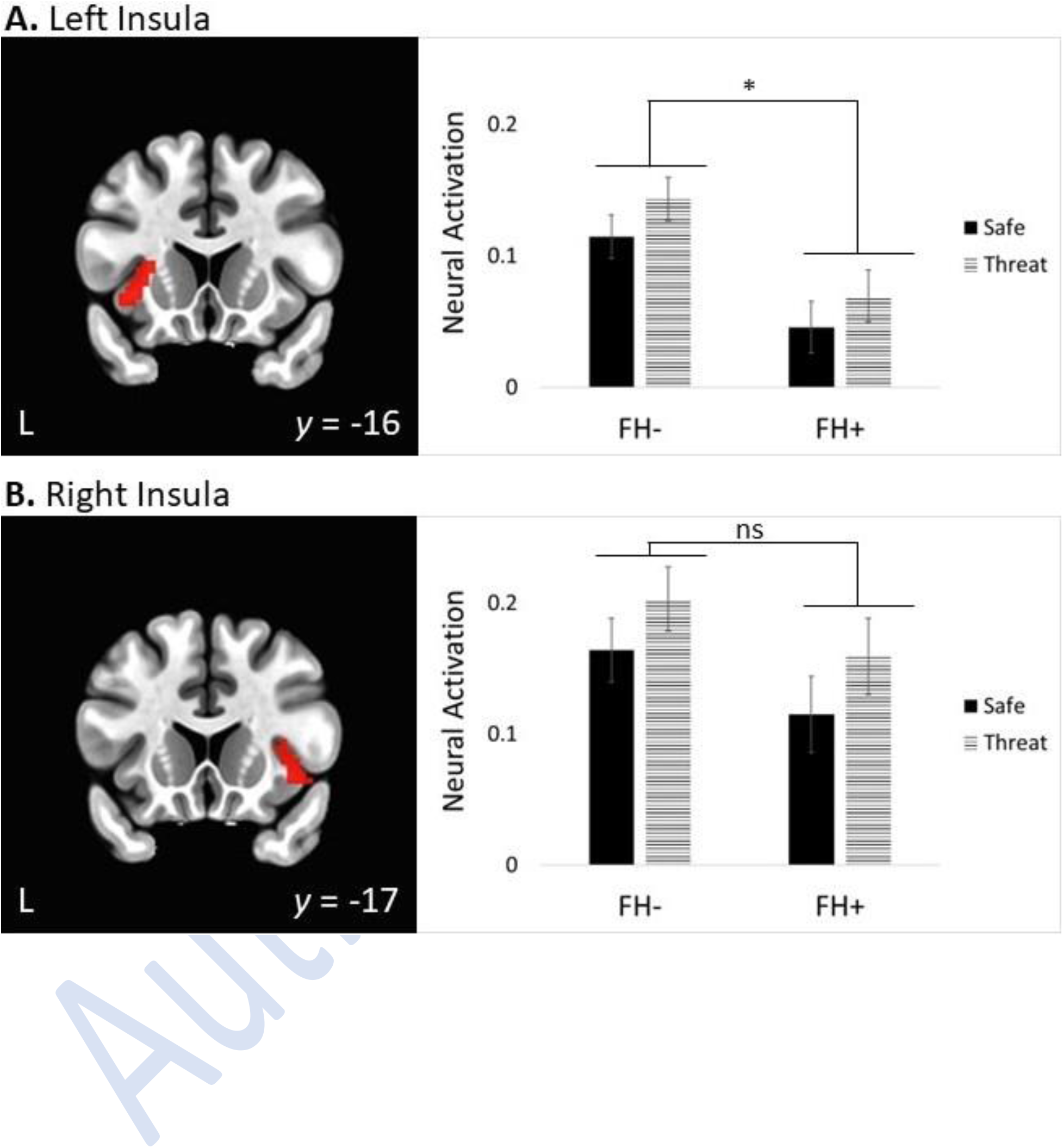
This figure depicts the results of the insula linear mixed effects modeling for the safe and threatening blocks during the MID-Scream Task. * indicates significance at *p* < .05, “ns” indicates non-significance, and all error bars represent the standard error of the least-squares means. FH-= family history negative and FH+ = family history positive. **Panel A:** The coronal view of the brain depicts the left insula highlighted in red. The graph depicts the main effects of group (*F*_1,73_ = 8.51, *p* = 0.005; indicated by the * on the graph) and block (*F*_1,73_ = 11.27, *p* = 0.001); the group-by-block interaction was not significant. **Panel B:** The coronal view of the brain depicts the right insula highlighted in red. The graph depicts the effects of group and block on neural activation; only block showed a significant main effect (*F*_2,365_ = 11.95, *p* = 0.001), while the main effect of group (indicated by the “ns” on the graph) and the group-by-block interaction were not significant.

**Table 2.**
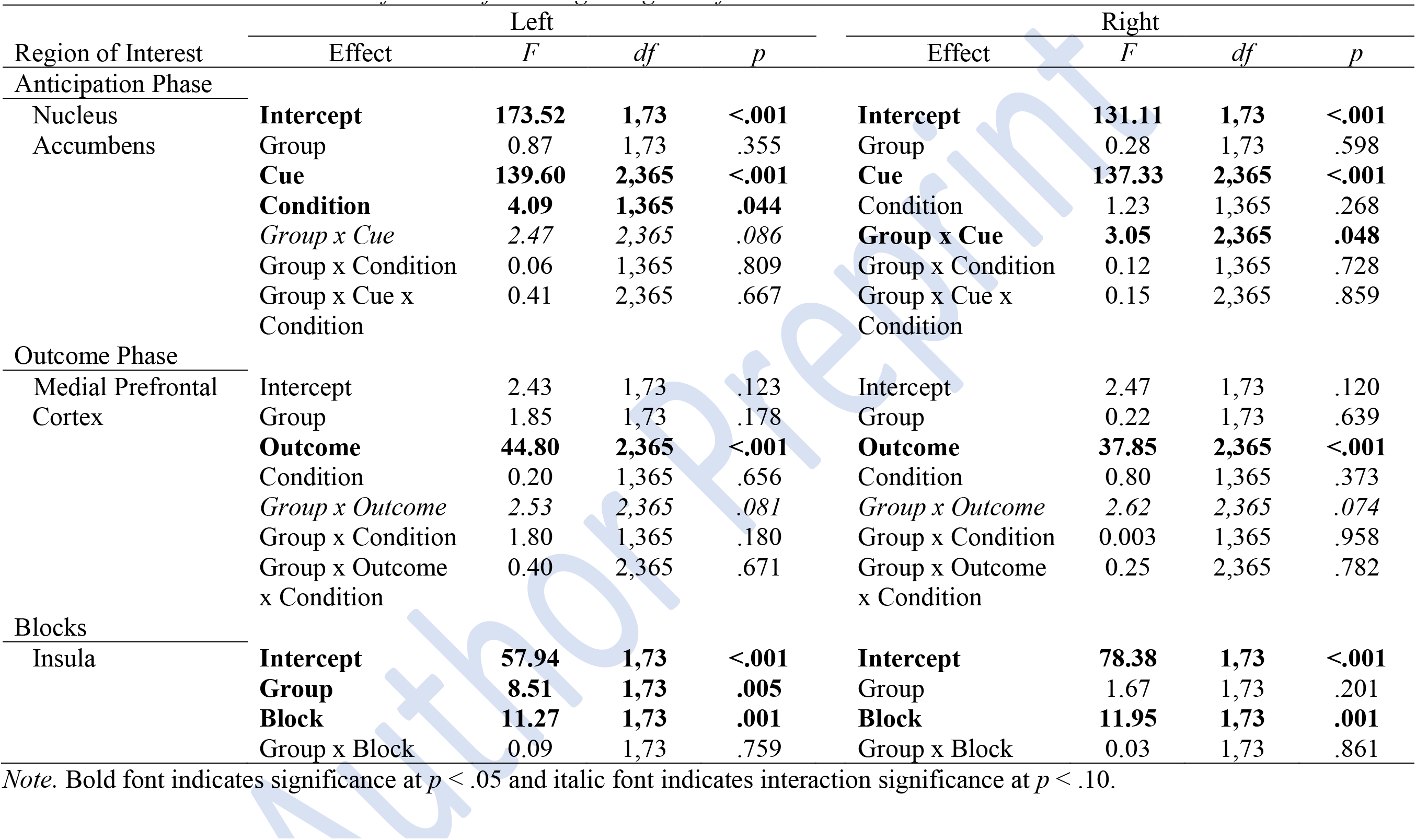
Factorial ANOVA Results for the Left and Right Regions of Interest.

#### Nucleus Accumbens

During the anticipation phase of the task in the left and right nucleus accumbens, there were no significant group-by-condition or group-by-cue-by-condition interactions (see Table 2). In the left accumbens only, there was a significant main effect of condition (*F*_1,365_ = 4.09, *p* = 0.044), where participants demonstrated greater activation during the safe relative to threat condition (Figure 4). Additionally, there was a significant main effect of cue (both *F*_2,365_ >137.0, *p* < 0.001) and a group-by-cue interaction (both *F*_2,365_ > 2.0, *p* < 0.09) on neural activation bilaterally in the accumbens (Figure 4). Participants demonstrated greater activation during cues to gain $5 relative to gain $0 and avoid losing $5, and during cues to avoid losing $5 relative to gain $0. The relative differences in activation between cues to gain $5 and gain $0 and between cues to avoid losing $5 and gain $0 were greater for the family history negative group compared to the family history positive group. The groups did not significantly differ on activation during cues to gain $5, avoid losing $5, or gain $0 (all *p* > 0.50).

**Figure 4.**
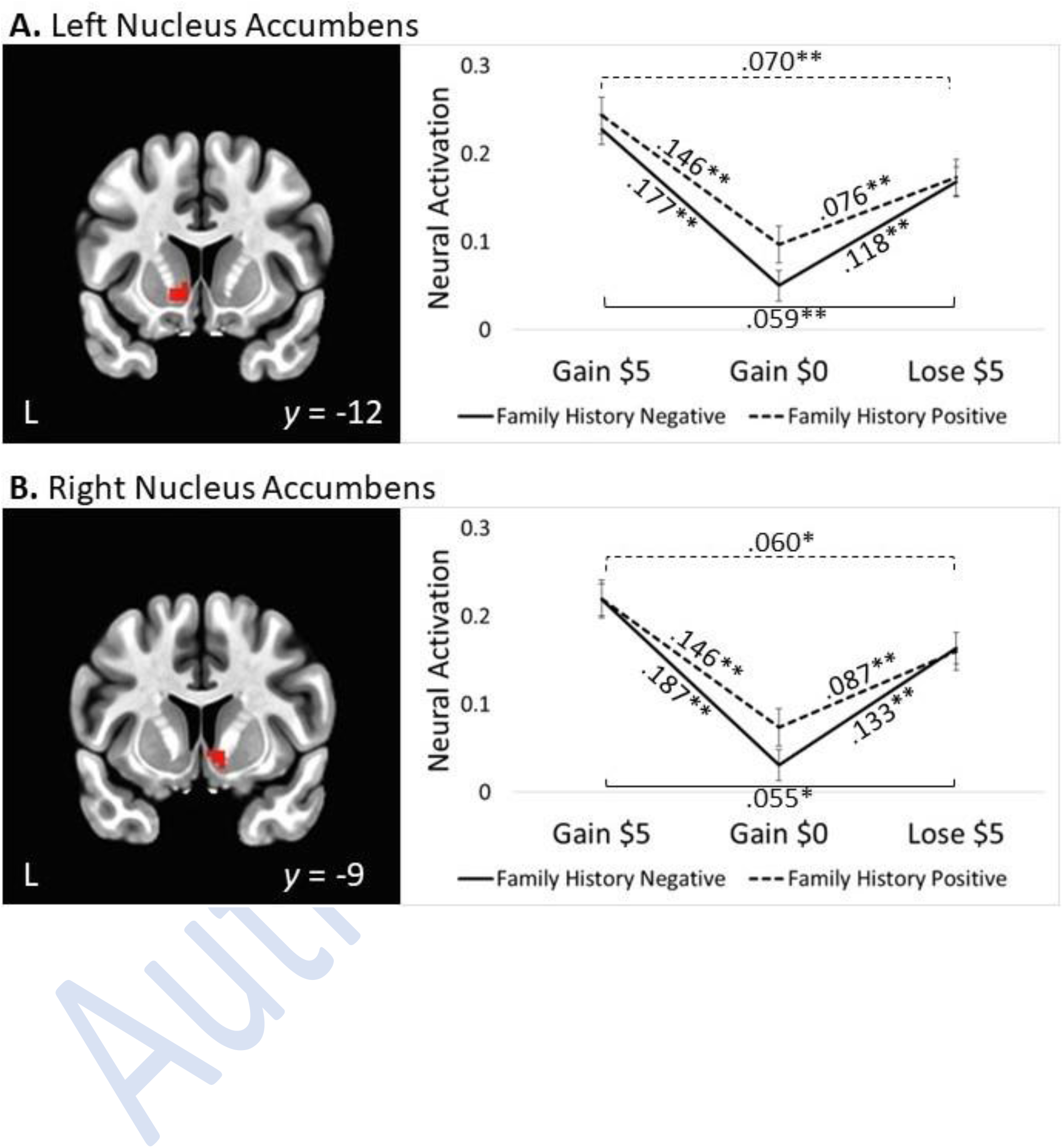
This figure depicts the results of the nucleus accumbens linear mixed effects modeling for the anticipation phase of the MID-Scream Task. Values on the graphs indicate the average relative difference in activation between cues, ** indicates significance at *p* < .001, and all error bars represent the standard error of the least-squares means. **Panel A:** The coronal view of the brain depicts the left nucleus accumbens highlighted in red. The graph depicts the significant group-by-cue interaction (*F*_2,365_ = 2.47, *p* = 0.086) with significant contrasts. **Panel B:** The coronal view of the brain depicts the right nucleus accumbens highlighted in red. The graph depicts the significant group-by-cue interaction (*F*_2,365_ = 3.05, *p* = 0.048) with significant contrasts.

#### Medial Prefrontal Cortex

During the outcome phase of the task, in the bilateral medial prefrontal cortex, there were no significant main effects of condition, nor significant group-by-condition or group-by-outcome-by-condition interactions (see Table 2). There was a significant main effect of outcome (both *F*_2,365_ > 37.0, *p* < 0.001) and a group-by-outcome interaction (both *F*_2,365_ > 2.0, *p* < 0.09) on activation bilaterally (Figure 5). Participants demonstrated greater activation during outcomes of winning $5 relative to gaining $0 and avoiding the loss of $5. The relative difference in activation between outcomes of winning $5 and gaining $0 was greater for the family history negative group bilaterally compared to the family history positive group. The groups did not significantly differ on activation during the winning $5, gaining $0, or the loss of $5 outcomes (all *p* > 0.10).

**Figure 5.**
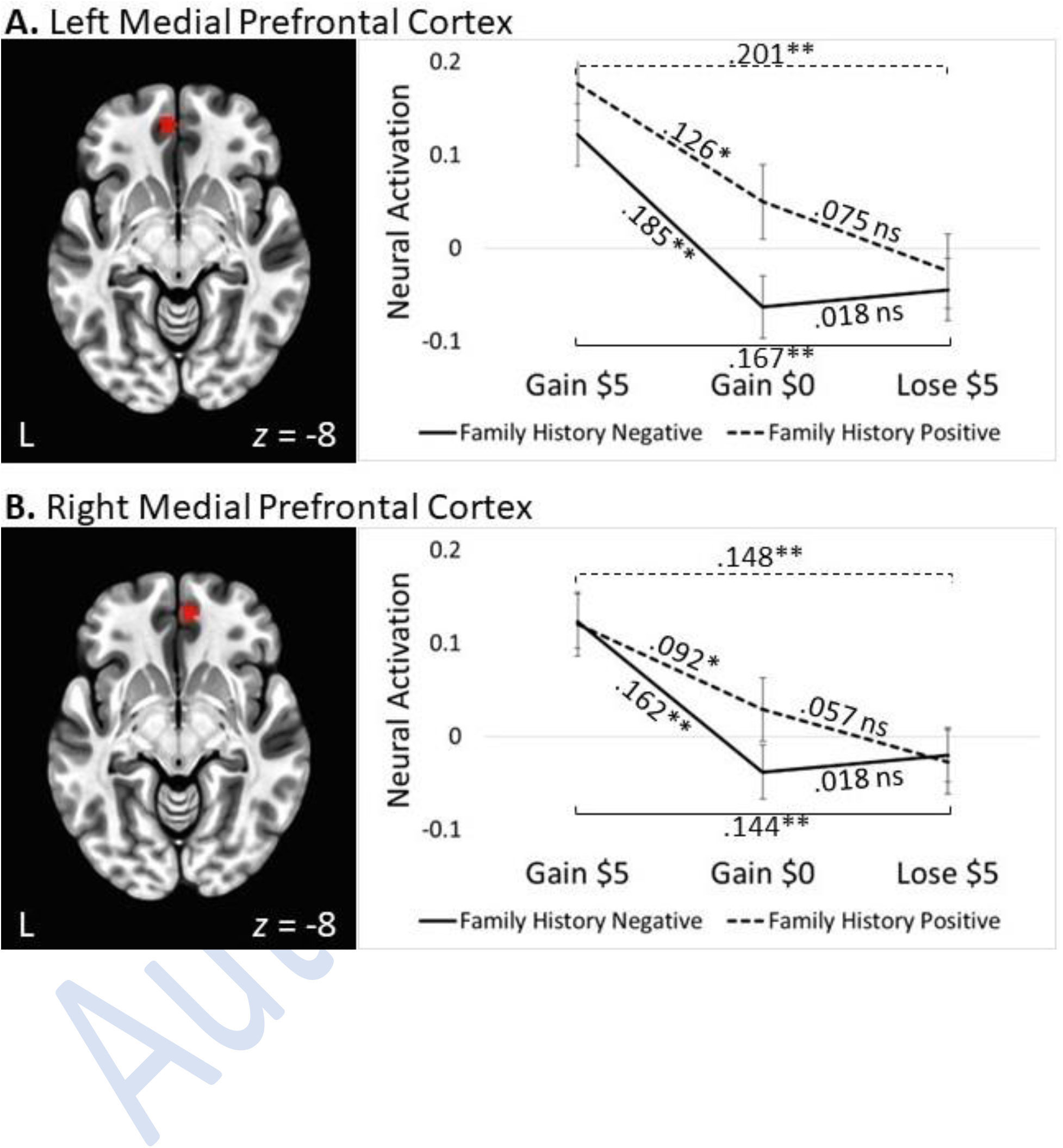
This figure depicts the results of the medial prefrontal cortex linear mixed effects modeling for the outcome phase of the MID-Scream Task. Lose $5 = avoiding losing $5 outcome, values on the graphs indicate the average relative difference in activation between outcomes, * indicates significance at *p* < .05, ** indicates significance at *p* < .001, “ns” indicates non-significance, and all error bars represent the standard error of the least-squares means. **Panel A:** The axial view of the brain depicts the left medial prefrontal cortex highlighted in red. The graph depicts the significant group-by-outcome interaction (*F*_2,365_ = 2.53, *p* = 0.081) with significant contrasts. **Panel B:** The axial view of the brain depicts the right medial prefrontal cortex highlighted in red. The graph depicts the significant group-by-outcome interaction (*F*_2,365_ = 2.62, *p* = 0.074) with significant contrasts.

#### Sensitivity Analyses

Results showed that the significant main and interaction effects in the unadjusted models were not altered after including lifetime alcohol use disorder, cannabis use disorder, sex, trait anxiety, or depression in the adjusted models. None of these variables had significant associations with neural activation in the bilateral insula, left nucleus accumbens, and bilateral medial prefrontal cortex (all *F*_1,69_ < 4.0, *p* ≥ .05), while trait anxiety was significantly associated with neural activation in the right nucleus accumbens (*F*_1,69_ = 4.86, *p* = .031); see Supporting Information.

### Exploratory Analyses

Whole-brain analyses did not identify any significant group-by-cue-by-condition effects. One cluster was identified with a significant effect of group and condition during the anticipation phase of the task. Regions in the cluster include the left operculum parietal 1, retroinsular cortex, and area PFcm of the inferior parietal lobule. In this cluster, there was a significant main effect of cue (*F*_2,365_ = 23.31, *p* < 0.001) and a significant group-by-condition interaction (*F*_1,365_ = 19.96, *p* < 0.001; Table S6 and Figure S2); see Supporting Information.

We explored effect size differences (41) to depict how the groups compared during the anticipation of threat the threatening blocks of the task, and also during the anticipation of winning money when presented with the gain $5 cue. During the threatening blocks, the family history negative relative to the family history positive group showed greater activation across several regions of the brain, with one significant cluster with a medium-large effect size located bilaterally in the dorsal anterior cingulate cortex, extending bilaterally to the caudal dorsomedial prefrontal cortex (*p* = 0.001, *d* = 0.75; Figure S3). The groups showed non-significant differences in activation in numerous areas during the anticipation of gaining $5, with some areas showing medium effect sizes (i.e., *d* > 0.5) for group differences (Figure S4); see Supporting Information.

## Discussion

The present study tested the hypothesis that healthy young adults with a family history of harmful alcohol use differ in reward processing while under sustained threat compared to those without such family history. Using the MID-Scream task (26), we examined the interactions between family history group, anticipation of unpredictable threat, and anticipation of monetary reward on neural activation in threat– and reward-related brain regions. We did not find support for the hypothesized interaction between family history, threat, and reward. Our results indicate that prior to the onset of an alcohol use disorder, individuals with enriched risk may not differ from those without such risk on sensitivity to threat but demonstrate diminished salience to potential rewarding and negative consequences.

Contrary to our hypothesis, the groups did not differ on neural activation in the insula as a function of induced anticipatory anxiety due to unpredictable threat, despite the family history positive group reporting higher levels of state– and trait-anxiety. This is contrary to previous research indicating that healthy individuals with higher levels of anxiety (42), and individuals with alcohol use disorder (10) show increased insular activation while under unpredictable threat. Further, there were no three-way group-by-cue-by-condition interactions in any of the ROIs. This may indicate that young adults with familial risk of developing an alcohol use disorder, but who have not yet transitioned to harmful alcohol use patterns, do not differ from those without such family histories in their sensitivity to threat. It may be that anticipation of threat may have a greater impact on reward processing should individuals go on to develop more harmful patterns of alcohol use (e.g., the substance-induced model; 43) in the future. Continued hazardous or harmful alcohol use is associated with the development of depression (44) and anxiety (43), and results in negative affectivity (45), and individuals may then continue to engage in harmful alcohol use due to the reduction of anxious states following the consumption of alcohol (4).

It is also possible that in our sample, those with enriched risk for developing an alcohol use disorder may not experience altered reactivity to prolonged threat due to resilience (19). It may be that by this age, if individuals with enriched risk have not gone on to develop harmful patterns of use, they may be more resilient to environmental risk factors including threatening events. However, should family history positive individuals go on to develop hazardous alcohol use behaviors in the future, we may expect to see altered insular functioning, given the association with alcohol craving (46), alcohol use disorder (10, 46), and risky decision making (47). Further, we may expect to see deficits in threat processing associated with reward anticipation and outcome via deactivation in the insula (21).

Consistent with previous research (e.g., 16, 19, 20), our results indicate that those with positive family histories have less neural differentiation between rewarding and neutral cues and outcomes, than those without such family histories. These results suggest that the family history positive group has dampened monetary reward salience irrespective of unpredictable threat. We found the family history positive group, versus the family history negative group, demonstrated smaller relative differences in neural activation in the nucleus accumbens and the medial prefrontal cortex during the anticipation and notification of winning money relative to earning no money, respectively. We further found that the family history positive group reported liking and being excited during the anticipation of winning money relative to winning no money, less so than the family history negative group. The family history positive group reported feeling less nervous during the anticipation of winning money relative to earning no money, compared to the family history negative group. These results are consistent with the findings of a recent meta-analysis (21) that found that individuals with alcohol use disorder demonstrated less neural activation in several reward and incentive brain regions, including the ventral striatum, during anticipation of reward and receipt of reward during the MID task. Zeng and colleagues (21) posit that decreased activation in these regions among individuals with alcohol use disorder indicate dysfunction in incentive salience to conventional rewards (e.g., money). Taken together, our results indicate that family history positive young adults may not be as sensitive to the rewarding salience of monetary reward tasks as family history negative individuals.

At the same time, negative outcomes, such as the actual loss of money does not appear to differentially alter reward response in family history positive individuals. This may indicate that healthy at-risk young adults may be appropriately sensitive to the experience of negative outcomes prior to the development of chronic heavy alcohol use. Meanwhile, the dampened response to the possibility of losing money relative to neutral outcomes and less self-reported nervousness during anticipation of winning or losing money among the family history positive group suggests dysfunction in the salience of *potential* negative consequences. Thus, it appears that family history positive individuals demonstrate decreased reward response to both potential rewarding and negative outcomes. The results of our study suggest that family history positive individuals have a predisposition to dysfunctional reward processing of potential conventional rewards and losses, even prior to engaging in hazardous alcohol use patterns. The lack of natural reward response has garnered speculation that such individuals may engage in excessive sensation seeking to achieve these rewarding effects, including using substances at hazardous levels (48).

### Limitations

Our study had some limitations. It is possible that our study was underpowered to detect three-way interactions. However, during exploratory analyses we followed rigorous false-positive correction, using a voxelwise threshold of *p* < 0.001 as recommended by Cox et al. (49), indicating that the lack of significant three-way interactions are feasible. The majority of individuals in our study were white, so the homogeneity of our sample may decrease generalizability. It is possible that our sample was resilient or comprised of individuals motivated to refrain from harmful drinking patterns. For example, our sample may demonstrate resilience as this sample represents a group of at-risk individuals who, despite having begun to drink alcohol, have not largely developed any hazardous drinking patterns, thus may have surpassed the window during which we would expect such young adults to develop harmful patterns of use. Finally, our study design did not counterbalance the order of presenting the threat and safe blocks; all participants experienced a threat block first. While we do not have reason to suspect this to have compromised the signal of the threat condition, we cannot rule out the possibility that it altered the response in some way.

### Conclusions

Through employing a model of both unpredictable, sustained threat and reward processing in a modified MID task, we studied an at-risk group for alcohol use disorder to address questions about neural processing prior to the development of problematic alcohol use patterns. The results did demonstrate important group differences based on family history. Most notably, the results of this study show that family history positive individuals did not differ from family history negative individuals in neural response in the insula during unpredictable threat, but did differ as a group by demonstrating diminished neural response to monetary incentives during the MID task in the nucleus accumbens and medial prefrontal cortex. These results may provide an important clue to how alcohol use disorder develops with regards to emotional processing. Our results suggest that at-risk individuals, who have not developed harmful alcohol use patterns, do demonstrate a diminished response to monetary reward, while notably not differing during a simulated anxious state. While this would require further study, our results may provide evidence that prior to developing harmful alcohol use patterns, there is a predisposition in enriched risk individuals through an altered reward pathway. With further study in more diverse samples, we may find that altered response to unpredictable threat may be a neural adaptation that follows from chronic, heavy alcohol use rather than a preexisting condition.

## Data Availability

All data produced in the present study are available upon reasonable request to the authors

## Acknowledgements

Study data were collected and managed using REDCap electronic data capture tools hosted at the University of Colorado. REDCap is a secure, web-based application designed to support data capture for research studies, providing: 1) an intuitive interface for validated data entry; 2) audit trials for tracking data manipulation and export procedures; 3) automated export procedures for seamless data downloads to common statistical packages; and 4) procedures for importing data from external sources.

