## Supplemental Materials for "NEURAL RESPONSE TO THREAT AND REWARD AMONG YOUNG ADULTS AT RISK FOR ALCOHOL USE DISORDER"

### Behavioral Outcomes of the MID-Scream Task

#### Reaction Time and Success Rate

There was no significant group-by-cue-by-condition interaction on reaction time or success rate during the MID-Scream task (see Table S1). For reaction time, there were significant main effects of cue and condition, and a significant group-by-cue effect (all $F > 4$, $p < 0.02$). Participants had significantly shorter reaction times during the gain $5 cue relative to the gain $0, and during the safe condition relative to threat. The family history negative group, but not the family history positive group, responded significantly faster to cues to gain $5 relative to gain $0; the other contrasts for both groups were non-significant (Figure S1). The groups did not significantly differ on reaction time during cues to gain $5, avoid losing $5, or gain $0 (all $p > 0.90$). For success rate, there was a significant main effect of cue ($F_{2,365} = 9.84$, $p < .001$). Participants were significantly more successful on both gain $5 and avoid losing $5 trials relative to gain $0 trials (Figure S1). There were no main effects of group or condition, or group-by-cue or group-by-condition interactions (see Table S1).

#### Self-Report Ratings

In addition to the results reported in the main text, we found significant main effects of cue on self-reported liking, excitement, and nervousness (all $F_{2,365} > 172.0$, $p < 0.001$; see Table S1). Participants reported liking cues to gain $5 significantly more than avoiding losing $5, and cues to gain $0 more than avoiding losing $5 (all $p > 0.001$). Participants reported being more excited during cues to gain $5 and avoid losing $5 than cues to gain $0, and cues to gain $5 more than cues to avoid losing $5 (all $p > 0.001$). Lastly, participants were more nervous during cues to gain $5 and avoid losing $5 relative to cues to gain $0, and more nervous during cues to avoid losing $5 relative to cues to gain $5 (all $p < 0.05$); see Figure 2 in the main text. For hearing the scream, while group differences were not statistically significant (all $t_{73} < 1.70$, $p > .10$), the family history positive group generally reported disliking the scream more than the family history negative group and tended to be more excited and nervous when hearing the scream than family history negative group (Table S2). Table S2 reports means, standard deviations, and group differences for reaction times and self-report ratings by family history group.

### Regions of Interest Analyses

#### Sensitivity Analyses

For the ROIs, we conducted sensitivity analyses to assess variables we suspected may impact our significant findings. As a history of alcohol or cannabis use disorders, trait anxiety, and depression may influence current findings, particularly for the family history positive group, and as sex was significantly different between the groups, we included these variables in the linear mixed effects models for the left and right insula, nucleus accumbens, and medial prefrontal cortex. The adjusted factorial ANOVAs revealed that the significant main and interaction effects in the unadjusted models remained significant, and the effects of history of substance use disorder, sex, and depression were not significant in any ROI (all $F_{1,69} < 4.0$, $p \geq$

Author Preprint

.05), while trait anxiety was significant in the right nucleus accumbens only ($F_{1,69}$ = 4.86, $p$ = .031). Results of the adjusted models are presented in Tables S3-S5.

### Exploratory Analysis

Whole-brain analyses identified one cluster (75 voxels, peak coordinates: $x$ = 46.5, $y$ = 43.5, $z$ = 25.5) that survived the minimum cluster size ($k \geq 49$) with a significant effect of group and condition on neural activation during the anticipation phase of the task. Regions in the cluster include the left operculum parietal 1, retroinsular cortex, and area PFcm of the inferior parietal lobule. In this cluster, there was a significant main effect of cue ($F_{2,365}$ = 23.31, $p$ < 0.001) and a significant group-by-condition interaction ($F_{1,365}$ = 19.96, $p$ < 0.001; Table S6 and Figure S2). Participants exhibited greater activation during cues to gain $5 and avoid losing $5 relative to gain $0, and during cues to gain $5 relative to avoid losing $5 (all $p$ < 0.004). Family history positive participants, but not family history negative showed significantly greater activation during the threat condition relative to the safe condition; the groups did not significantly differ during the threat or safe conditions (both $p$ > 0.50). Table S6 presents the factorial ANOVA results of the linear mixed effects model for the identified cluster with a significant effect of group and condition on neural activation, during the exploratory whole-brain analyses. Figure S2 shows a sagittal view of the brain where the significant cluster is highlighted in shades of yellow and interaction results are depicted in the graphs.

As we expected group to have wider effect on neural activation across the brain during the MID-Scream task, we examined the magnitude of the effect of family history group during anticipation of gaining $5 when presented with the gain $5 cue, and also during sustained threat during the threatening blocks of the task. Effect size maps using Cohen's $d$ are presented in Figures S3 and S4.

Author Preprint

**Table S1.** *Factorial ANOVA Results for Reaction Time, Success Rate, and Self-Report Ratings on MID-Scream fMRI Task*

| Reaction Time & Success Rate | | | |
|---|---|---|---|
| Effect | *F* | *df* | *p* |
| Reaction Time | | | |
| **Intercept** | **8442.66** | **1,73** | **<.001** |
| Group | 0.09 | 1,73 | .766 |
| **Cue** | **6.27** | **2,365** | **.002** |
| **Condition** | **31.69** | **1,365** | **<.001** |
| **Group x Cue** | **4.19** | **2,365** | **.016** |
| Group x Condition | 1.96 | 1,365 | .163 |
| Group x Cue x Condition | 0.09 | 2,365 | .917 |
| Success Rate | | | |
| **Intercept** | **8025.36** | **1,73** | **<.001** |
| Group | 1.73 | 1,73 | .193 |
| **Cue** | **9.84** | **2,365** | **<.001** |
| Condition | 0.01 | 1,365 | .924 |
| Group x Cue | 0.22 | 2,365 | .804 |
| Group x Condition | 0.37 | 1,365 | .544 |
| Group x Cue x Condition | 1.32 | 2,365 | .268 |

| Self-Report Ratings | | | |
|---|---|---|---|
| Effect | *F* | *df* | *p* |
| Liking | | | |
| **Intercept** | **1970.28** | **1,73** | **<.001** |
| Group | 0.11 | 1,73 | .740 |
| **Cue** | **302.39** | **2,365** | **<.001** |
| **Condition** | **26.99** | **1,365** | **<.001** |
| **Group x Cue** | **6.86** | **2,365** | **.001** |
| Group x Condition | 0.22 | 1,365 | .637 |
| Group x Cue x Condition | 0.15 | 2,365 | .864 |
| Excitement | | | |
| **Intercept** | **1872.53** | **1,73** | **<.001** |
| Group | 1.79 | 1,73 | .185 |
| **Cue** | **226.71** | **2,365** | **<.001** |
| Condition | 0.12 | 1,365 | .733 |
| **Group x Cue** | **8.93** | **2,365** | **<.001** |
| Group x Condition | 1.25 | 1,365 | .265 |
| Group x Cue x Condition | 0.04 | 2,365 | .962 |
| Nervousness | | | |
| **Intercept** | **1135.02** | **1,73** | **<.001** |
| Group | 2.02 | 1,73 | .160 |
| **Cue** | **172.91** | **2,365** | **<.001** |
| **Condition** | **69.57** | **1,365** | **<.001** |
| *Group x Cue* | *2.83* | *2,365* | *.060* |
| Group x Condition | 1.46 | 1,365 | .228 |
| Group x Cue x Condition | 0.03 | 2,365 | .969 |

*Note.* Bold font indicates significance at $p < .05$ and italic font indicates interaction significance at $p < .10$.

Author Preprint

**Figure S1. Reaction Time and Success Rate.** This figure depicts the results of the reaction time and success rate linear mixed effects modeling. Values indicate the average relative difference in reaction times and success rates, **indicates significance at $p < 0.001$, "ns" indicates $p > 0.05$, and all error bars represent the standard error of the least-squares means. **Panel A:** The first graph depicts the group-by-cue interaction for reaction times. Family history negative responded significantly faster during cues to potentially win money relative to earning no money. While the main effect of condition was significant, the group-by-condition interaction was non-significant (second graph). Overall, participants had faster reaction times to the safe condition relative to the threat condition. **Panel B:** While the main effect of cue was significant, the two graphs depict the non-significant group-by-cue and group-by-condition interactions, respectively. Overall, participants were more successful after seeing cues to potentially win money or avoid losing money relative to gaining no money.

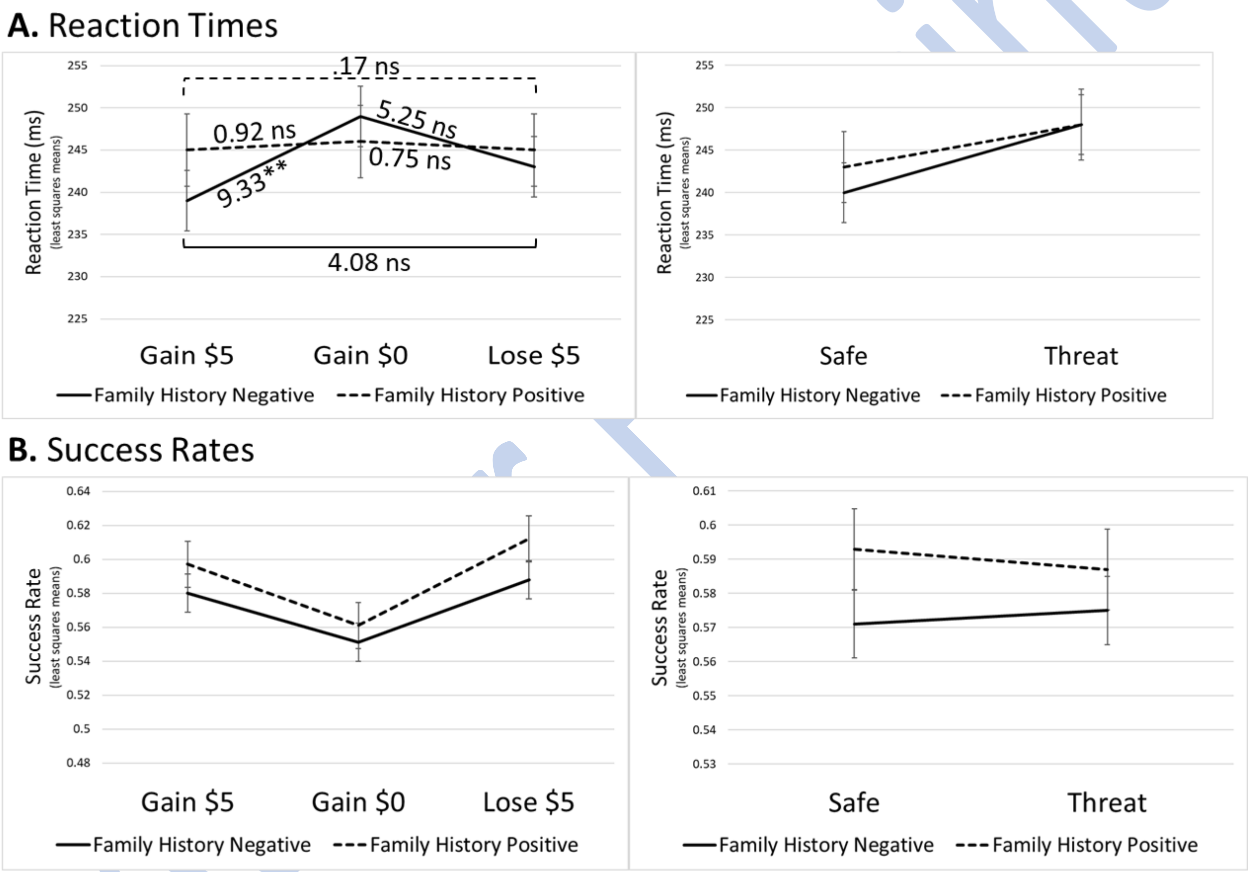

**Table S2.** *Mean Reaction Times and Self-Report Ratings by Group for Cue and Condition Trials*

| Variable | FH- (*n* = 44) *M* (*SD*) | FH+ (*n* = 31) *M* (*SD*) | Test Statistic |
|---|---|---|---|
| Reaction Times | | | |
| Gain \$5 - Safe | 236.11 (29.33) | 242.87 (24.35) | *t*(73) = 1.05 |
| Gain \$5 - Threat | 242.58(24.53) | 247.22(25.31) | *t*(73) = 0.80 |
| Lose \$5 - Safe | 236.98(25.98) | 240.43(21.82) | *t*(73) = 0.60 |
| Lose \$5 - Threat | 249.87 (26.78) | 250 (21.45) | *t*(73) = 0.02 |
| Gain \$0 - Safe | 245.84 (26.73) | 245.39 (24.02) | *t*(73) = 0.07 |
| Gain \$0 - Threat | 251.51 (25.54) | 246.53 (24.51) | *t*(73) = 0.85 |
| Self-reported Liking | | | |
| **Gain \$5 - Safe** | **9.25 (1.3)** | **8.29 (1.83)** | ***t*(50.57) = 2.51** |
| Gain \$5 - Threat | 8 (2.17) | 7.1(2.3) | *t*(73) = 1.73 |
| Lose \$5 - Safe | 1.93 (2.25) | 2.94 (2.32) | *t*(73) = 1.88 |
| Lose \$5 - Threat | 1.75 (2.43) | 2.29 (2.34) | *t*(73) = 0.96 |
| Gain \$0 - Safe | 5.98 (2.17) | 6.45 (2.16) | *t*(73) = 0.93 |
| Gain \$0 - Threat | 4.61 (2.09) | 4.94 (2.17) | *t*(73) = 0.65 |
| Self-reported Excitement | | | |
| **Gain \$5 - Safe** | **9.02 (1.13)** | **7.71 (2.37)** | ***t*(39.69) = 2.87** |
| **Gain \$5 - Threat** | **8.61 (1.42)** | **7.68 (1.76)** | ***t*(73) = 2.55** |
| Lose \$5 - Safe | 6.73 (1.83) | 5.9 (1.68) | *t*(73) = 1.98 |
| Lose \$5 - Threat | 6.57 (2.08) | 6.23 (2.09) | *t*(73) = 0.70 |
| Gain \$0 - Safe | 3.73 (2.18) | 4.16 (2.46) | *t*(73) = 0.80 |
| Gain \$0 - Threat | 3.57 (2.36) | 4.26 (2.42) | *t*(73) = 1.23 |
| Self-reported Nervousness | | | |
| Gain \$5 - Safe | 5.95 (2.72) | 5.84 (1.93) | *t*(73) = 0.20 |
| Gain \$5 - Threat | 7.05 (2.44) | 7.26 (1.48) | *t*(71.67) = 0.47 |
| Lose \$5 - Safe | 6.82 (2.24) | 6.84 (1.97) | *t*(73) = 0.04 |
| Lose \$5 - Threat | 7.3 (2.09) | 7.87 (1.38) | *t*(72.79) = 1.44 |
| Gain \$0 - Safe | 1.5 (1.7) | 2.35 (2.51) | *t*(49.05) = 1.65 |
| Gain \$0 - Threat | 3.91 (2.97) | 5.23 (2.89) | *t*(73) = 1.91 |
| Hearing the Scream | | | |
| Liking | 2.86 (2.51) | 2.00 (1.84) | *t*(73) = 1.63 |
| Excitement | 5.52 (3.37) | 6.13 (2.66) | *t*(73) = 0.84 |
| Nervousness | 6.09 (3.27) | 7.03 (3.05) | *t*(73) = 1.26 |

*Note.* FH+ = family history positive, FH- = family history negative.
Bold font indicates significance at $p < .05$.

Author Preprint

**Table S3.** *Sensitivity Analyses: Adjusted Factorial ANOVA Results for the Left and Right Insula for the Safe and Threatening Blocks of the MID-Scream Task*

| | Left | | | | Right | | | |
|---|---|---|---|---|---|---|---|---|
| Region of Interest | Effect | *F* | *df* | *p* | Effect | *F* | *df* | *p* |
| Insula | Intercept | 1.87 | 1,69 | .176 | Intercept | 2.37 | 1,69 | .128 |
| | **Group** | **5.34** | **1,69** | **.024** | Group | 1.09 | 1,69 | .300 |
| | **Block** | **11.27** | **1,73** | **.001** | **Block** | **11.95** | **1,73** | **.001** |
| | SUD History | 0.03 | 1,69 | .853 | SUD History | 0.27 | 1,69 | .606 |
| | Sex | 0.003 | 1,69 | .960 | Sex | 0.05 | 1,69 | .825 |
| | Trait Anxiety | 0.03 | 1,69 | .865 | Trait Anxiety | 0.04 | 1,69 | .850 |
| | Depression | 0.68 | 1,69 | .412 | Depression | 0.07 | 1,69 | .792 |
| | Group x Block | 0.09 | 1,73 | .759 | Group x Block | 0.03 | 1,73 | .861 |

*Note.* Bold font indicates significance at $p < .05$. SUD = substance use disorder.

**Table S4.** *Sensitivity Analyses: Adjusted Factorial ANOVA Results for the Left and Right Nucleus Accumbens for the Anticipation Phase of the MID-Scream Task*

| | Left | | | | Right | | | |
|---|---|---|---|---|---|---|---|---|
| Region of Interest | Effect | *F* | *df* | *p* | Effect | *F* | *df* | *p* |
| Nucleus Accumbens | **Intercept** | **16.04** | **1,69** | **<.001** | **Intercept** | **17.06** | **1,69** | **<.001** |
| | Group | 1.51 | 1,69 | .223 | Group | 0.77 | 1,69 | .385 |
| | **Cue** | **139.60** | **2,365** | **<.001** | **Cue** | **137.33** | **2,365** | **<.001** |
| | **Condition** | **4.09** | **1,365** | **.044** | Condition | 1.23 | 1,365 | .268 |
| | SUD History | 0.48 | 1,69 | .491 | SUD History | 0.001 | 1,69 | .979 |
| | Sex | 0.80 | 1,69 | .374 | Sex | 1.33 | 1,69 | .254 |
| | Trait Anxiety | 3.97 | 1,69 | .050 | **Trait Anxiety** | **4.86** | **1,69** | **.031** |
| | Depression | 3.06 | 1,69 | .086 | Depression | 2.97 | 1,69 | .090 |
| | *Group x Cue* | *2.47* | *2,365* | *.086* | **Group x Cue** | **3.05** | **2,365** | **.048** |
| | Group x Condition | 0.06 | 1,365 | .809 | Group x Condition | 0.12 | 1,365 | .728 |
| | Group x Cue x Condition | 0.41 | 2,365 | .667 | Group x Cue x Condition | 0.15 | 2,365 | .859 |

*Note.* Bold font indicates significance at $p < .05$ and italic font indicates interaction significance at $p < .10$. SUD = substance use disorder.

Author Preprint

**Table S5.** *Sensitivity Analyses: Adjusted Factorial ANOVA Results for the Left and Right Medial Prefrontal Cortex for the Outcome Phase of the MID-Scream Task*

| | Left | | | | Right | | | |
|---|---|---|---|---|---|---|---|---|
| Region of Interest | Effect | *F* | *df* | *p* | Effect | *F* | *df* | *p* |
| Medial Prefrontal Cortex | Intercept | 0.47 | 1,69 | .496 | Intercept | 1.37 | 1,69 | .248 |
| | Group | .935 | 1,69 | .337 | Group | 0.06 | 1,69 | .804 |
| | **Outcome** | **44.80** | **2,365** | **<.001** | **Outcome** | **37.85** | **2,365** | **<.001** |
| | Condition | 0.20 | 1,365 | .656 | Condition | 0.80 | 1,365 | .373 |
| | SUD History | 0.05 | 1,69 | .822 | SUD History | 0.04 | 1,69 | .859 |
| | Sex | 0.20 | 1,69 | .655 | Sex | 0.04 | 1,69 | .852 |
| | Trait Anxiety | 1.49 | 1,69 | .227 | Trait Anxiety | 2.37 | 1,69 | .128 |
| | Depression | 2.36 | 1,69 | .129 | Depression | 1.46 | 1,69 | .232 |
| | *Group x Outcome* | *2.53* | *2,365* | *.081* | *Group x Outcome* | *2.62* | *2,365* | *.074* |
| | Group x Condition | 1.80 | 1,365 | .181 | Group x Condition | 0.003 | 1,365 | .958 |
| | Group x Outcome x Condition | 0.40 | 2,365 | .671 | Group x Outcome x Condition | 0.25 | 2,365 | .782 |

*Note*. Bold font indicates significance at $p < .05$ and italic font indicates interaction significance at $p < .10$. SUD = substance use disorder.

Author Preprint

**Table S6.** *Exploratory Analysis: Factorial ANOVA Results for Significant Effect of Group and Condition on Activation During Whole-Brain Analysis at k ≥ 49 (p ≤ .001)*

| Effect | *F* | *df* | *p* |
|---|---|---|---|
| **Intercept** | **21.76** | **1,73** | **<.001** |
| Group | 0.12 | 1,73 | .734 |
| **Cue** | **23.31** | **2,365** | **<.001** |
| Condition | 2.84 | 1,365 | .093 |
| Group x Cue | 0.39 | 2,365 | .679 |
| **Group x Condition** | **19.96** | **1,365** | **<.001** |
| Group x Cue x Condition | 1.48 | 2,365 | .230 |

*Note.* Bold font indicates significance at $p < .05$.

Preprint

**Figure S2. Whole-Brain Analysis: Effect of Group and Condition on Neural Activation.** The figure below depicts the significant linear mixed effects results of the exploratory whole-brain analyses during which a cluster of 75 voxels located in the left operculum parietal 1, retroinsular cortex, and area PFcm of the inferior parietal lobule (depicted in the sagittal view and highlighted in yellow) survived the significance threshold of $k \geq 49$, $p < .001$. Within this cluster, there was a significant main effect of cue and a significant group-by-condition effect on neural activation during the task. The first graph depicts the non-significant group-by-cue interaction, and the second graph depicts the significant group-by-condition interaction with one significant contrast. *Note:* Values on the graph indicate the average relative difference in activation between conditions, ** indicates significance at $p < .001$, "ns" indicates non-significance, and all error bars represent the standard error of the least-squares means.

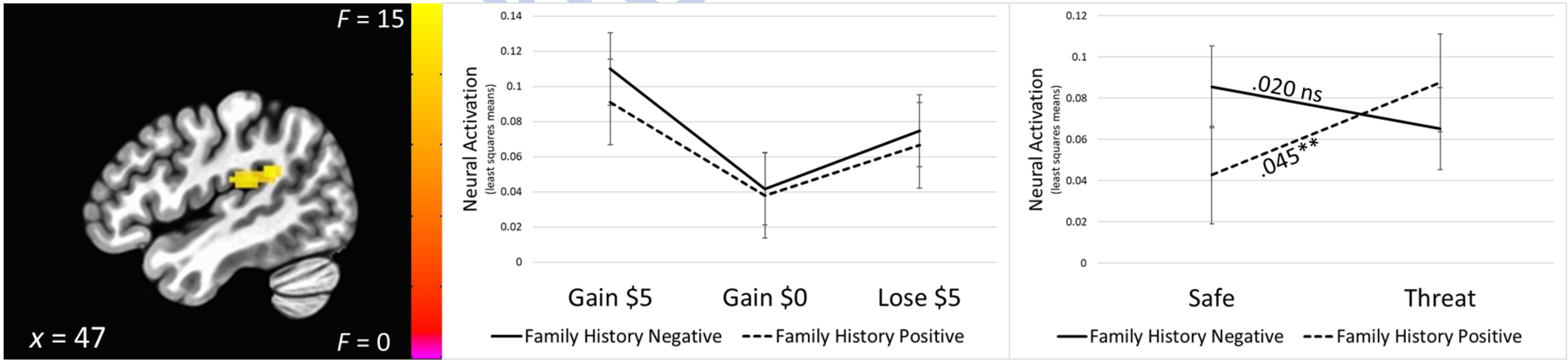

**Figure S3. Effect Size Map: Group Comparison During Anticipation of Winning (Gain $5).** The figure below depicts effect size (Cohen's *d*) differences between the family history negative and family history positive groups during the anticipation of winning money (the gain $5 cue). Warmer colors (e.g., oranges) indicate brain regions where the family history negative group demonstrated greater neural activation during the gain $5 cue, while cooler colors (e.g., blues) indicate brain regions where the family history positive group demonstrated greater activation during the gain $5 cue. As can be seen, the groups demonstrated differences in neural activation in several areas including prefrontal, medial, and visual regions. However, these differences did not meet the minimum voxel-wise threshold of $k \geq 49$, $p < 0.001$. Effect sizes are generally small across these regions. **Panel A:** The image displays a montage in the axial view. **Panel B:** The top image depicts the coronal view and shows very little differential activation in the accumbens during the anticipation of winning money, and the bottom image depicts the sagittal view.

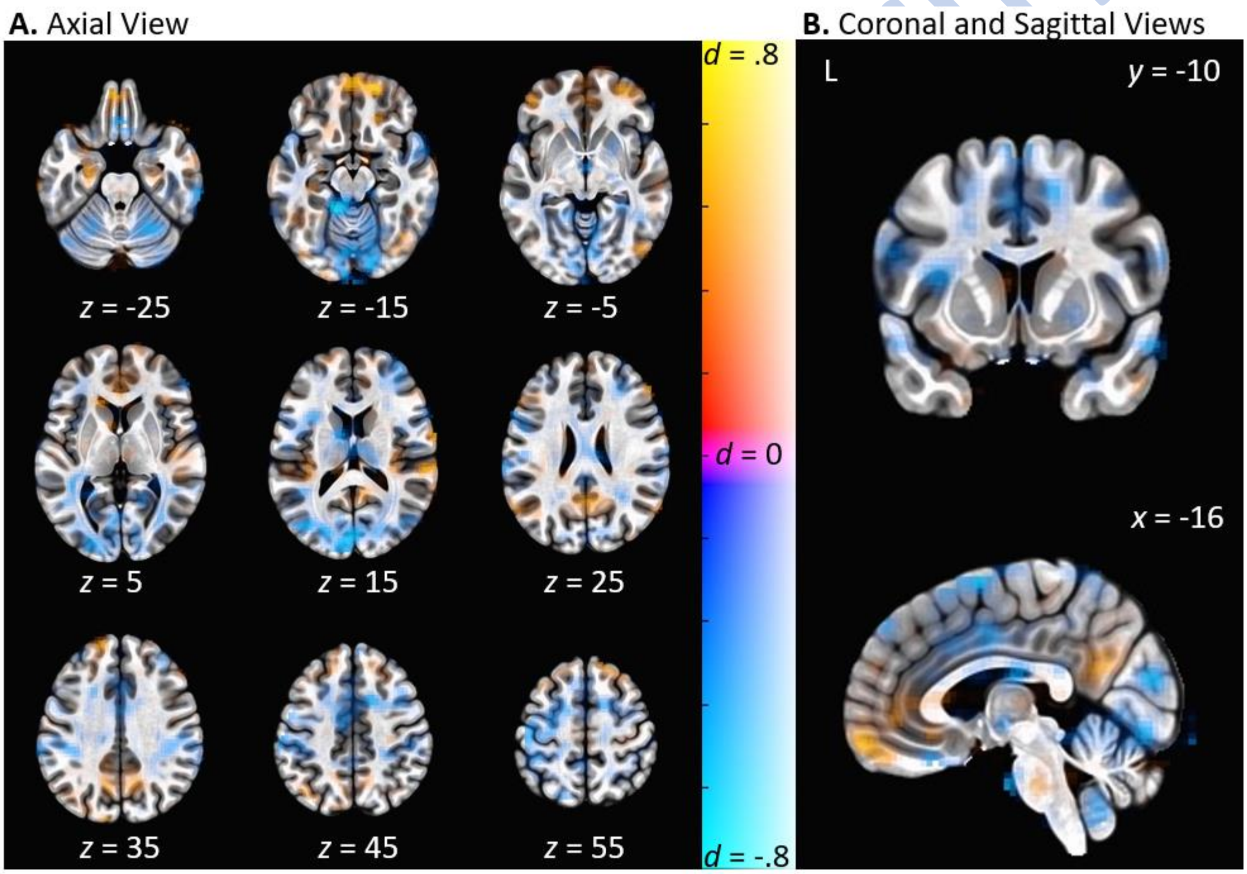

**Figure S4. Effect Size Map: Group Comparison During Threat Blocks.** The figure below depicts effect size (Cohen's *d*) differences between the family history negative and family history positive groups during the threat block of the task. Warmer colors (e.g., oranges) indicate brain regions where the family history negative group demonstrated greater neural activation during the threat block, while cooler colors (e.g., blues) indicate brain regions where the family history positive group demonstrated greater activation during the threat block. The black outlines on colored regions indicate the significant cluster of activation (62 voxels). As can be seen, the family history negative group demonstrated greater neural activation in several areas, with one significant cluster surviving the minimum voxel-wise threshold of $k \geq 49$, $p < 0.001$, with a medium-large effect size located bilaterally in the dorsal anterior cingulate cortex, extending bilaterally to the caudal dorsomedial prefrontal cortex ($t = 3.41$, $d = 0.75$). Effect sizes are generally medium across the other brain regions. **Panel A:** The image displays a montage in the axial view. **Panel B:** The top image depicts the coronal view, and the bottom image depicts the sagittal view.

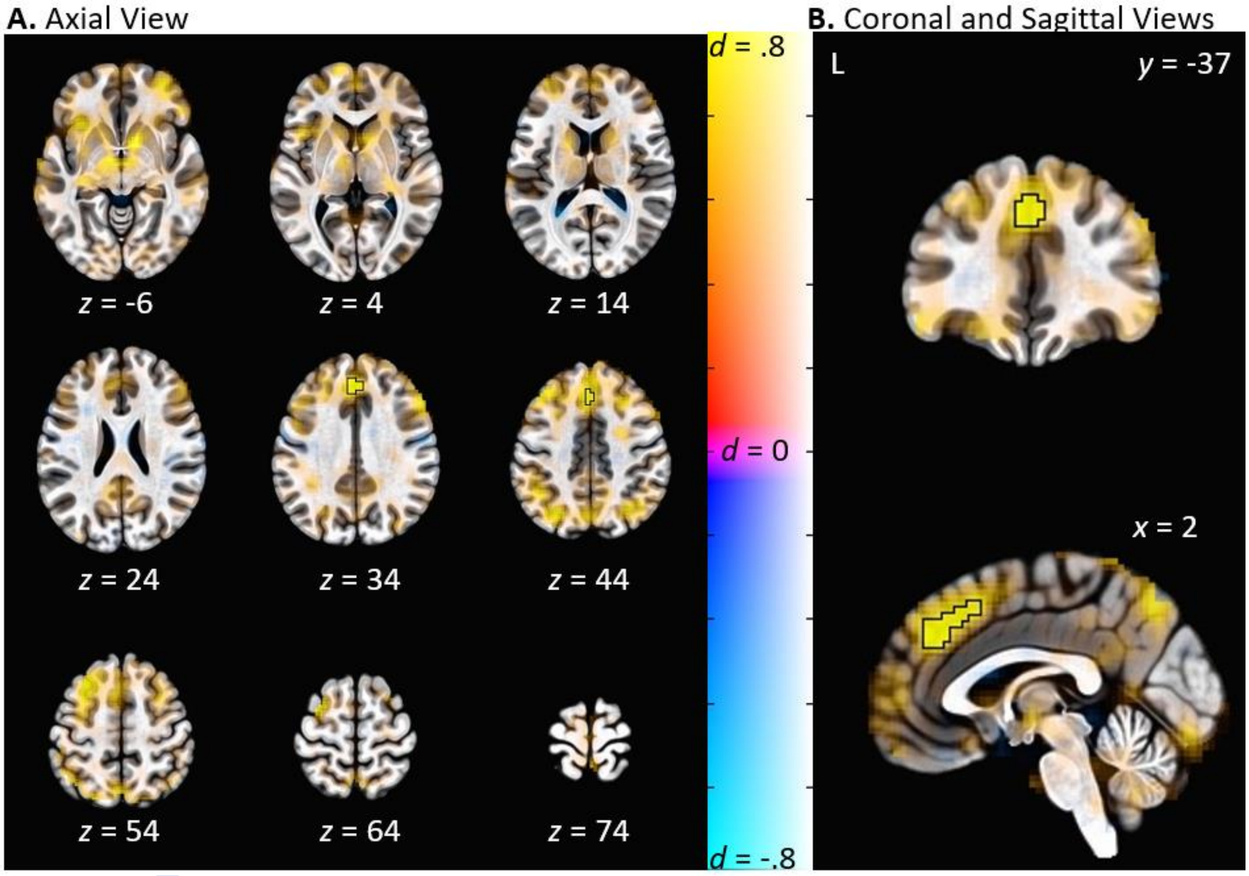
